# Syndemic burden, menopause, and cardiovascular risk among midlife and older women with and without HIV

**DOI:** 10.64898/2026.11.03.26367225

**Authors:** Yue Pan, Emily M. Cherenack, Daniel J. Feaster, Yawen Lu, BreAnne Young, Kayla V. McNeely, Tracey Wilson, Brandilyn A. Peters, Emily B. Levitan, Caitlin Moran, Catalina Ramirez, Elizabeth F. Topper, Kathleen Weber, Rebecca Abelman, Amanda B. Spence, Candice A. Sternberg, Nicholas Fonseca Nogueira, Claudia A. Martinez, Maria L. Alcaide, Deborah L. Jones

## Abstract

Midlife and older women living with HIV often carry several co-occurring social and behavioral stressors, and how that accumulated burden relates to cardiovascular risk is not well described. We examined whether cumulative syndemic burden was associated with estimated 10-year risk of atherosclerotic cardiovascular disease among women aged 45 years and older, and whether menopause stage modified it. Data came from the MACS/WIHS Combined Cohort Study, a multisite United States cohort, for visits between October 2020 and March 2025. The full sample included 1,993 women, 1,446 of them living with HIV; a secondary sample of 1,063 women virally suppressed at baseline contributed 4,009 visits. Syndemic burden was the sum of six conditions: elevated depressive symptoms, current tobacco use, hazardous alcohol use, recent stimulant use, physical inactivity, and material insecurity. Ten-year cardiovascular risk was estimated with the Pooled Cohort Equations and modeled with linear mixed-effects models with random intercepts; random forest classifiers were trained separately to predict high estimated risk (7.5 percent or greater). Median age was 56 years, 63 percent were non-Hispanic Black, and 57 percent reported two or more syndemic conditions. Each additional syndemic condition was associated with a 0.40-point higher estimated risk score (95 percent confidence interval 0.01 to 0.78; p=0.043), and postmenopausal status with a 1.3-point higher score than premenopause (0.39 to 2.3; p=0.006). The syndemic burden by menopause stage interaction was not significant overall (p=0.2), although an exploratory signal was present among virally suppressed women (0.50; p=0.079). The classifiers discriminated moderately (areas under the curve 0.727 and 0.704); serum creatinine, body mass index, syndemic burden, and postmenopausal status ranked among the most important predictors. Syndemic burden and reproductive aging were each independently and modestly associated with higher estimated cardiovascular risk, supporting joint assessment of both in risk stratification for midlife women with and without HIV.

## Introduction

Cardiovascular disease (CVD) is the leading cause of death among women in the United States and remains a dominant driver of morbidity across the life course [1]. Among people living with HIV (PLWH), improved survival with contemporary antiretroviral therapy has shifted clinical priorities toward chronic comorbidities, including CVD [2]. An American Heart Association scientific statement emphasizes that CVD risk in PLWH reflects the combined influence of traditional cardiometabolic risk factors and HIV-related mechanisms, including persistent immune activation and systemic inflammation [3]. Among women living with HIV (WLWH), cardiovascular vulnerability is multifactorial, involving intersecting pathways linking traditional risk factors, metabolic dysregulation, reproductive aging, and chronic immune activation [4]. Studying this vulnerability requires analytic strategies that incorporate reproductive aging stage and contextual risk domains alongside traditional clinical factors.

Syndemic theory provides a framework for integrating these contextual drivers. Syndemics describe the clustering of multiple adverse conditions within harmful social contexts, producing excess disease burden beyond what any single condition would predict [5]. In HIV research, syndemic frameworks conceptualize how co-occurring psychosocial and behavioral conditions concentrate in marginalized populations and jointly shape health vulnerability [6]. Empirically, syndemic patterns involving unhealthy alcohol use, smoking, and depressive symptoms have been associated with increased incident CVD among people with and without HIV, supporting the relevance of cumulative burden to cardiovascular risk stratification [7].

Menopause represents a biologically distinct interval of reproductive aging during which endocrine remodeling intersects with cardiometabolic physiology. Longitudinal and mechanistic evidence demonstrated that the menopause transition is associated with distinct patterns of change in endogenous sex hormones and adverse shifts in body fat distribution, lipids and lipoproteins, and vascular health, processes that are plausibly relevant for subsequent ASCVD risk and prevention timing [8].

Standard cardiovascular risk prediction tools - including the ACC/AHA Pooled Cohort Equations (PCE) [9] - have demonstrated systematic underestimation of observed CVD risk in HIV cohorts, with calibration problems particularly pronounced in women [10,11]. PCE does not incorporate psychosocial and structural determinants of cardiovascular health, including material insecurity, depression, and substance use, which are especially prevalent among WLWH and may independently drive cardiometabolic risk trajectories [12–16].

Despite this rationale, much prior CVD research in HIV has emphasized isolated risk factors rather than cumulative burden of co-occurring vulnerabilities. This leaves a gap in the literature for WLWH, for whom depression, tobacco and substance use, physical inactivity, and material hardship may cluster within individuals and potentially amplify cardiovascular vulnerability through behavioral and psychosocial mechanisms. A syndemic approach is therefore well suited to characterize how multiple co-occurring conditions relate to cardiovascular risk profiles and cardiovascular morbidity in WLWH, while recognizing that such measures may reflect broader vulnerability rather than a direct causal mechanism. Using longitudinal data from the MACS/WIHS Combined Cohort Study, a large observational cohort designed to understand chronic comorbidities among PLWH and comparable without HIV participants [17], this study evaluates syndemic burden and menopause stage as primary exposures in relation to estimated 10-year ASCVD risk (continuous and dichotomous at ≥7.5%), and uses random forest classifiers to characterize important predictors of high ASCVD risk in a population with intersecting biological and psychosocial risk domains.

## Materials and methods Ethics statement

This study is a secondary analysis of de-identified data from the MACS/WIHS Combined Cohort Study (MWCCS). The MWCCS protocol was approved by the institutional review board at each participating site, all participants provided written informed consent, and the study followed the Declaration of Helsinki. The present analysis was conducted under an approved MWCCS data analysis request. Data were accessed for research purposes on 20 October 2025, and the authors did not have access to information that could identify individual participants during or after data collection.

## Study design and population

MWCCS is a prospective observational cohort in the United States designed to study chronic health conditions affecting PLWH and comparable participants without HIV, integrating legacy cohorts and harmonized follow-up. Details have been previously described [17]. For the present longitudinal analysis, baseline and follow-up data were obtained through MWCCS visit windows spanning V101 (Oct 1, 2020-Sep 30, 2021), V102 (Oct 1, 2021-Sep 30, 2022), V103 (Oct 1, 2022-Sep 30, 2023), V104 (Oct 1, 2023-Sep 30, 2024), and V1051 (Oct 1, 2024-Mar 31, 2025).

Participants living with or without HIV were eligible for the full analytic cohort if they were women enrolled in MWCCS, were aged ≥45 years, and contributed at least two study visits during V101-V1051. Participant flow and analytic sample construction are summarized in Fig 1. Two analytic populations were prespecified. The primary population was the full analytic sample comprising 1,993 women, including 1,446 women living with HIV (WLWH) and 547 women without HIV (Fig 1). The secondary analytic population was restricted to WLWH who were virally suppressed at baseline (HIV RNA <50 copies/mL), yielding 1,063 participants; 383 WLWH were excluded from this sub-sample because they were not suppressed at baseline or had missing baseline viral load. The suppressed WLWH sub-sample contributed 4,009 total visits across follow-up.

**Fig 1.**
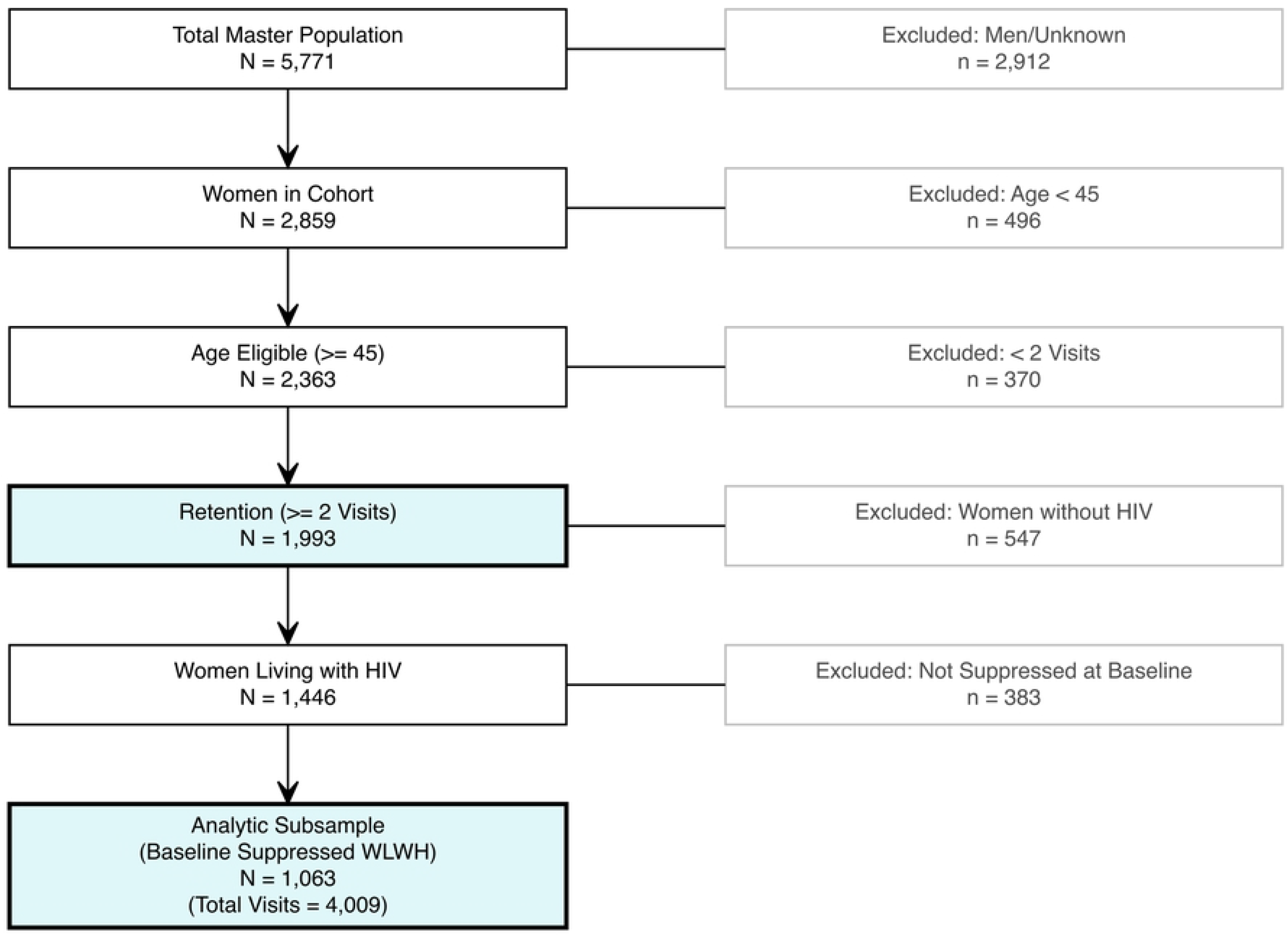
Participant flow diagram for the study sample. Participant selection from the MACS/WIHS Combined Cohort Study (MWCCS). Women aged 45 years or older seen between October 2020 and March 2025 were eligible. The full analytic sample comprised 1,993 women (1,446 women living with HIV [WLWH]; 547 women without HIV). The virally suppressed sub- cohort included 1,063 WLWH contributing 4,009 person-visits.

## Measures Primary outcomes

The primary continuous cardiovascular outcome was the 10-year ASCVD risk score, modeled longitudinally as a continuous measure. ASCVD risk estimation followed the ACC/AHA pooled cohort equations (PCE) framework [9], which uses age, race, lipids, blood pressure and treatment, diabetes status, and smoking status to estimate 10-year ASCVD risk. A binary high- risk outcome was also defined as 10-year ASCVD risk ≥7.5%, reflecting a commonly used guideline-relevant threshold for risk stratification and preventive therapy discussions.

## Primary exposures

Syndemic burden was operationalized as a cumulative syndemic score ranging from 0 to 6, derived from six binary conditions defined in the analytic dataset: elevated depressive symptoms, current tobacco use, hazardous alcohol use, recent stimulant use, physical inactivity, and material insecurity. For descriptive analyses, syndemic burden was also categorized as 0 conditions, 1 condition, and ≥2 conditions.

Menopause stage was modeled as a categorical exposure aligned with reproductive aging staging principles and the STRAW+10 framework, operationalized as premenopause, perimenopause, and postmenopause (natural and/or surgical).

## Covariates

Models adjusted for sociodemographic and clinical covariates including education, employment, insurance, BMI, hypertension, diabetes, and dyslipidemia. HIV-related factors were incorporated when appropriate. Age and race/ethnicity were excluded from ASCVD risk models to avoid circular adjustment, as these variables are embedded in the PCE.

The random forest models used 32 predictors spanning behavioral/syndemic factors, clinical comorbidities, biomarkers, psychosocial variables, and cognitive indicators, with PCE-embedded variables intentionally excluded to avoid tautological prediction.

## Statistical analysis

Descriptive statistics were used to summarize baseline demographic, clinical, and behavioral characteristics of the participants, stratified by the HIV status and syndemic factors among WLWH, and also for syndemic factors among virally suppressed WLWH subsample.

Particularly, we used median (Q1, Q3) for continuous variables and n (%) for categorical variables, with group comparisons using Kruskal-Wallis rank-sum tests for continuous variables and Fisher’s exact tests for categorical variables.

Linear mixed-effects models with random intercepts assessed longitudinal associations with continuous ASCVD risk. Random slopes were not included given convergence considerations. Models included time, syndemic score, menopause stage, and their interaction. HIV status was a fixed effect in the full sample and omitted by design in the suppressed sub-sample.

Complementing the mixed-effects regression approach, random forest classifiers were used to predict high ASCVD risk (≥7.5%), without pre-specifying functional form or interaction terms, thereby capturing potential nonlinear relationships and complex interactions among predictors. Random forests are ensemble tree methods that aggregate predictions across many decorrelated decision trees to reduce variance and improve generalization. Model development followed a participant-level train-test workflow, such that all visits from a given participant were assigned to either the training set or the held-out test set to avoid within-person leakage. The training/test split, stratified by outcome, was 70/30 for the full analytic sample and 60/40 for the virally suppressed sub-cohort. With the number of trees fixed at 1,000, hyperparameters (number of predictors sampled at each split and minimum node size) were tuned using 5-fold cross- validation on the training set, and performance was evaluated on the held-out test set using AUC, sensitivity, specificity, PPV, and NPV. Variable importance was assessed using impurity-based (Gini) importance; these rankings are descriptive and should not be interpreted as causal estimates. Models were developed separately for the full analytic sample and the virally suppressed sub-cohort. Reporting followed TRIPOD recommendations for clinical prediction models [18]. Missing data were addressed using multiple imputation by chained equations (MICE; m=5 imputations, maxit=5 iterations), with logistic regression for binary variables, predictive mean matching for continuous variables, and polytomous regression for nominal variables. All analyses were performed in R (version 4.5.2).

## Results

### Descriptive statistics for the full analytic sample and virally suppressed sub-sample

Participant eligibility criteria for the analysis are summarized in Fig 1. From the master MWCCS population (N=5,771), men and participants of unknown sex were excluded (n=2,912), yielding 2,859 women; women aged <45 were excluded (n=496), yielding 2,363 age-eligible women; participants with <2 visits were excluded (n=370), yielding the retained full analytic sample of 1,993 women with ≥2 visits. Among WLWH in the retained analytic cohort (n=1,446), those not virally suppressed at baseline (or missing baseline viral load) were excluded (n=383), yielding the baseline virally suppressed WLWH sub-sample of 1,063 participants with 4,009 total visits (Fig 1).

Baseline characteristics for the full analytic sample by HIV status are presented in Table 1. The full analytic cohort included 1,993 women (547 without HIV and 1,446 WLWH), with a median age of 56 years (Q1, Q3: 50, 61) and no evidence of age differences by HIV status (p=0.5; Table 1). Race/ethnicity distribution differed by HIV status (p=0.003; Table 1), with the overall cohort comprised of 63% non-Hispanic Black (1,265/1,993), 14% Hispanic (281/1,993), 8.2% non- Hispanic White (163/1,993), and 14% non-Hispanic other/multi (284/1,993). Cardiometabolic comorbidity prevalence was high, including hypertension in 73% (n=1,374) and diabetes in 21% (n=393), with dyslipidemia modestly more common in WLWH (69%) than women without HIV (64%) (p=0.039; Table 1). Median baseline 10-year ASCVD risk was 5 (2, 11) overall and was slightly higher among women without HIV at 6 (2, 12) compared with WLWH at 5 (2, 10) (p=0.028; Table 1). Syndemic burden was common, with 57% of women classified as having ≥2 syndemic conditions (1,133/1,993), 27% with 1 condition (536/1,993), and 16% with no syndemic conditions (324/1,993), and the 0/1/≥2 syndemic burden category distribution did not differ significantly by HIV status (p=0.2; Table 1).

**Table 1.**
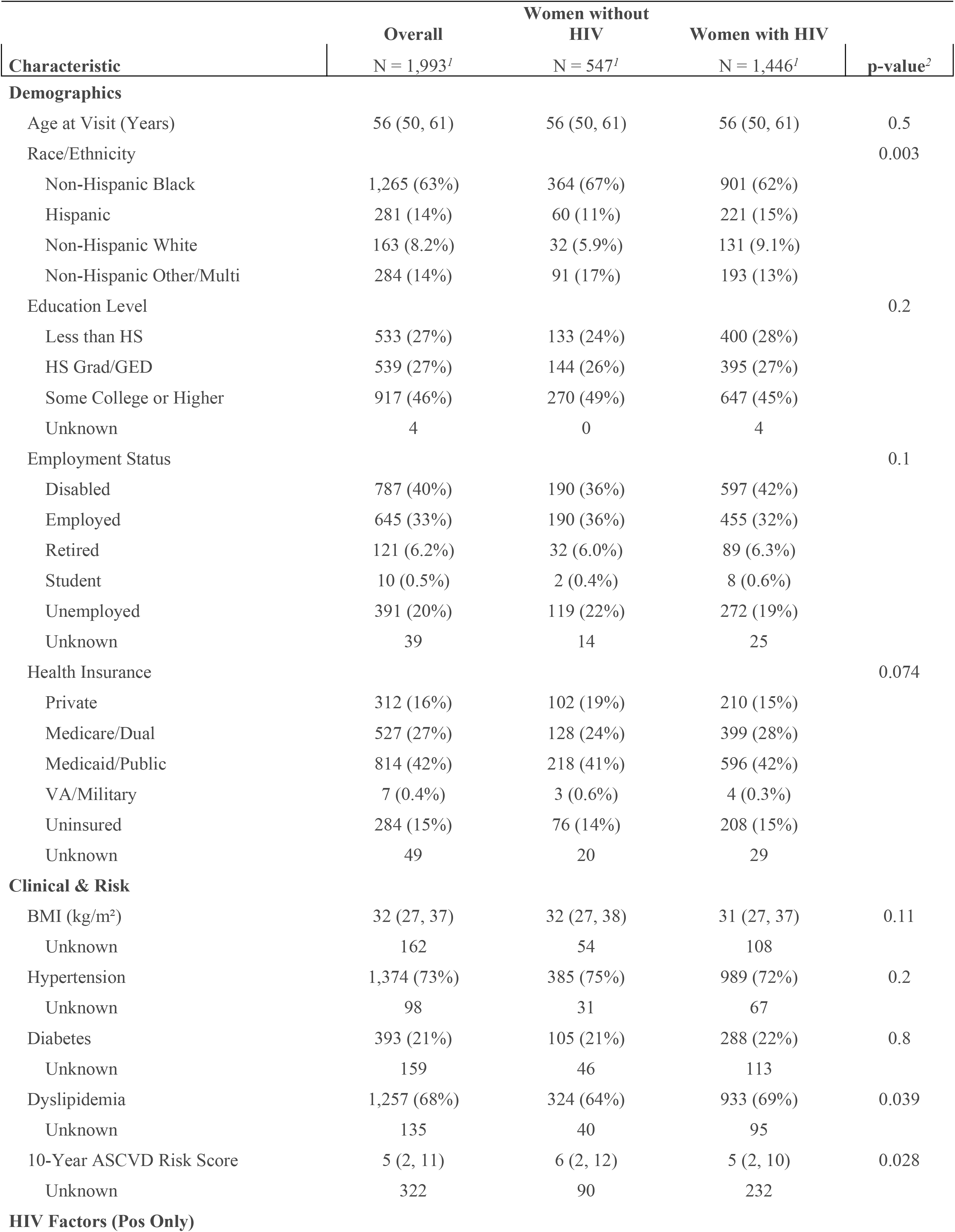
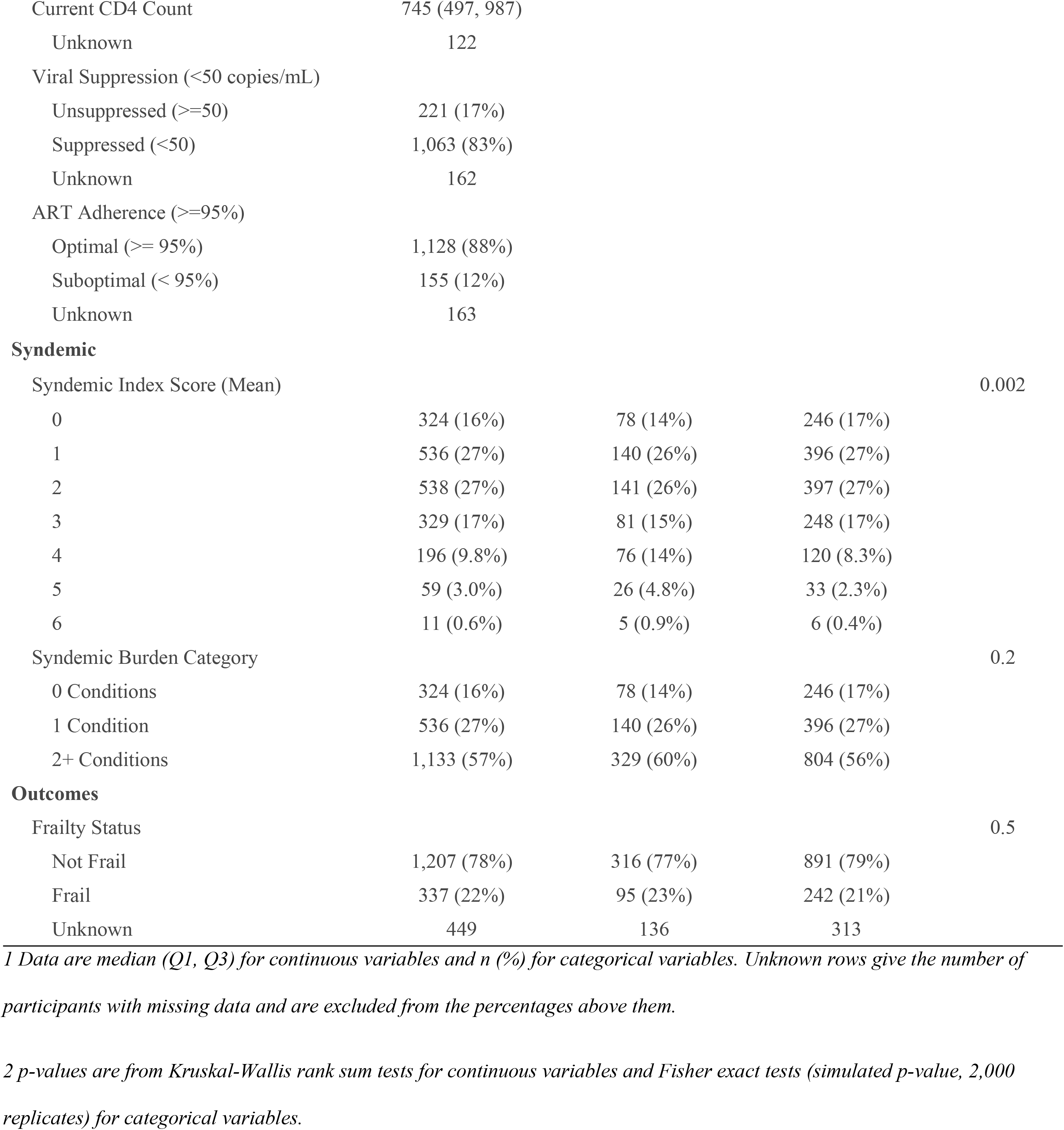
Full sample characteristics by HIV status.

|  | **Overall** | **Women without HIV** | **Women with HIV** |  |
|---|---|---|---|---|
| **Characteristic** | N = 1,993<sup>*1*</sup> | N = 547<sup>*1*</sup> | N = 1,446<sup>*1*</sup> | **p-value**<sup>*2*</sup> |
| **Demographics** |  |  |  |  |
| Age at Visit (Years) | 56 (50, 61) | 56 (50, 61) | 56 (50, 61) | 0.5 |
| Race/Ethnicity |  |  |  | 0.003 |
| Non-Hispanic Black | 1,265 (63%) | 364 (67%) | 901 (62%) |  |
| Hispanic | 281 (14%) | 60 (11%) | 221 (15%) |  |
| Non-Hispanic White | 163 (8.2%) | 32 (5.9%) | 131 (9.1%) |  |
| Non-Hispanic Other/Multi | 284 (14%) | 91 (17%) | 193 (13%) |  |
| Education Level |  |  |  | 0.2 |
| Less than HS | 533 (27%) | 133 (24%) | 400 (28%) |  |
| HS Grad/GED | 539 (27%) | 144 (26%) | 395 (27%) |  |
| Some College or Higher | 917 (46%) | 270 (49%) | 647 (45%) |  |
| Unknown | 4 | 0 | 4 |  |
| Employment Status |  |  |  | 0.1 |
| Disabled | 787 (40%) | 190 (36%) | 597 (42%) |  |
| Employed | 645 (33%) | 190 (36%) | 455 (32%) |  |
| Retired | 121 (6.2%) | 32 (6.0%) | 89 (6.3%) |  |
| Student | 10 (0.5%) | 2 (0.4%) | 8 (0.6%) |  |
| Unemployed | 391 (20%) | 119 (22%) | 272 (19%) |  |
| Unknown | 39 | 14 | 25 |  |
| Health Insurance |  |  |  | 0.074 |
| Private | 312 (16%) | 102 (19%) | 210 (15%) |  |
| Medicare/Dual | 527 (27%) | 128 (24%) | 399 (28%) |  |
| Medicaid/Public | 814 (42%) | 218 (41%) | 596 (42%) |  |
| VA/Military | 7 (0.4%) | 3 (0.6%) | 4 (0.3%) |  |
| Uninsured | 284 (15%) | 76 (14%) | 208 (15%) |  |
| Unknown | 49 | 20 | 29 |  |
| **Clinical & Risk** |  |  |  |  |
| BMI (kg/m²) | 32 (27, 37) | 32 (27, 38) | 31 (27, 37) | 0.11 |
| Unknown | 162 | 54 | 108 |  |
| Hypertension | 1,374 (73%) | 385 (75%) | 989 (72%) | 0.2 |
| Unknown | 98 | 31 | 67 |  |
| Diabetes | 393 (21%) | 105 (21%) | 288 (22%) | 0.8 |
| Unknown | 159 | 46 | 113 |  |
| Dyslipidemia | 1,257 (68%) | 324 (64%) | 933 (69%) | 0.039 |
| Unknown | 135 | 40 | 95 |  |
| 10-Year ASCVD Risk Score | 5 (2, 11) | 6 (2, 12) | 5 (2, 10) | 0.028 |
| Unknown | 322 | 90 | 232 |  |
| **HIV Factors (Pos Only)** |  |  |  |  |
| Current CD4 Count | 745 (497, 987) |  |  |  |
| Unknown | 122 |  |  |  |
| Viral Suppression (<50 copies/mL) |  |  |  |  |
| Unsuppressed (>=50) | 221 (17%) |  |  |  |
| Suppressed (<50) | 1,063 (83%) |  |  |  |
| Unknown | 162 |  |  |  |
| ART Adherence (>=95%) |  |  |  |  |
| Optimal (>= 95%) | 1,128 (88%) |  |  |  |
| Suboptimal (< 95%) | 155 (12%) |  |  |  |
| Unknown | 163 |  |  |  |
| **Syndemic** |  |  |  |  |
| Syndemic Index Score (Mean) |  |  |  | 0.002 |
| 0 | 324 (16%) | 78 (14%) | 246 (17%) |  |
| 1 | 536 (27%) | 140 (26%) | 396 (27%) |  |
| 2 | 538 (27%) | 141 (26%) | 397 (27%) |  |
| 3 | 329 (17%) | 81 (15%) | 248 (17%) |  |
| 4 | 196 (9.8%) | 76 (14%) | 120 (8.3%) |  |
| 5 | 59 (3.0%) | 26 (4.8%) | 33 (2.3%) |  |
| 6 | 11 (0.6%) | 5 (0.9%) | 6 (0.4%) |  |
| Syndemic Burden Category |  |  |  | 0.2 |
| 0 Conditions | 324 (16%) | 78 (14%) | 246 (17%) |  |
| 1 Condition | 536 (27%) | 140 (26%) | 396 (27%) |  |
| 2+ Conditions | 1,133 (57%) | 329 (60%) | 804 (56%) |  |
| **Outcomes** |  |  |  |  |
| Frailty Status |  |  |  | 0.5 |
| Not Frail | 1,207 (78%) | 316 (77%) | 891 (79%) |  |
| Frail | 337 (22%) | 95 (23%) | 242 (21%) |  |
| Unknown | 449 | 136 | 313 |  |

*1 Data are median (Q1, Q3) for continuous variables and n (%) for categorical variables. Unknown rows give the number of participants with missing data and are excluded from the percentages above them.*
*2 p-values are from Kruskal-Wallis rank sum tests for continuous variables and Fisher exact tests (simulated p-value, 2,000 replicates) for categorical variables.*

Full-sample baseline characteristics by syndemic burden are shown in Table 2. Women with higher syndemic burden differed across multiple sociodemographic indicators, including education (p<0.001), employment/disability (p<0.001), and insurance status (p<0.001; Table 2). Compared with women with 0 syndemic conditions, those with ≥2 conditions had substantially higher disability prevalence (54% vs 12%) and markedly lower private insurance coverage (6.1% vs 40%; Table 2). Median baseline ASCVD risk increased stepwise with syndemic burden, from 3 (1, 6) in those with 0 conditions to 4 (2, 10) in those with 1 condition and 6 (3, 12) in those with ≥2 conditions (p<0.001; Table 2). Frailty prevalence increased across syndemic strata, with frail status in 7.2% among women with 0 conditions and 30% among women with ≥2 conditions (p<0.001; Table 2).

**Table 2.**
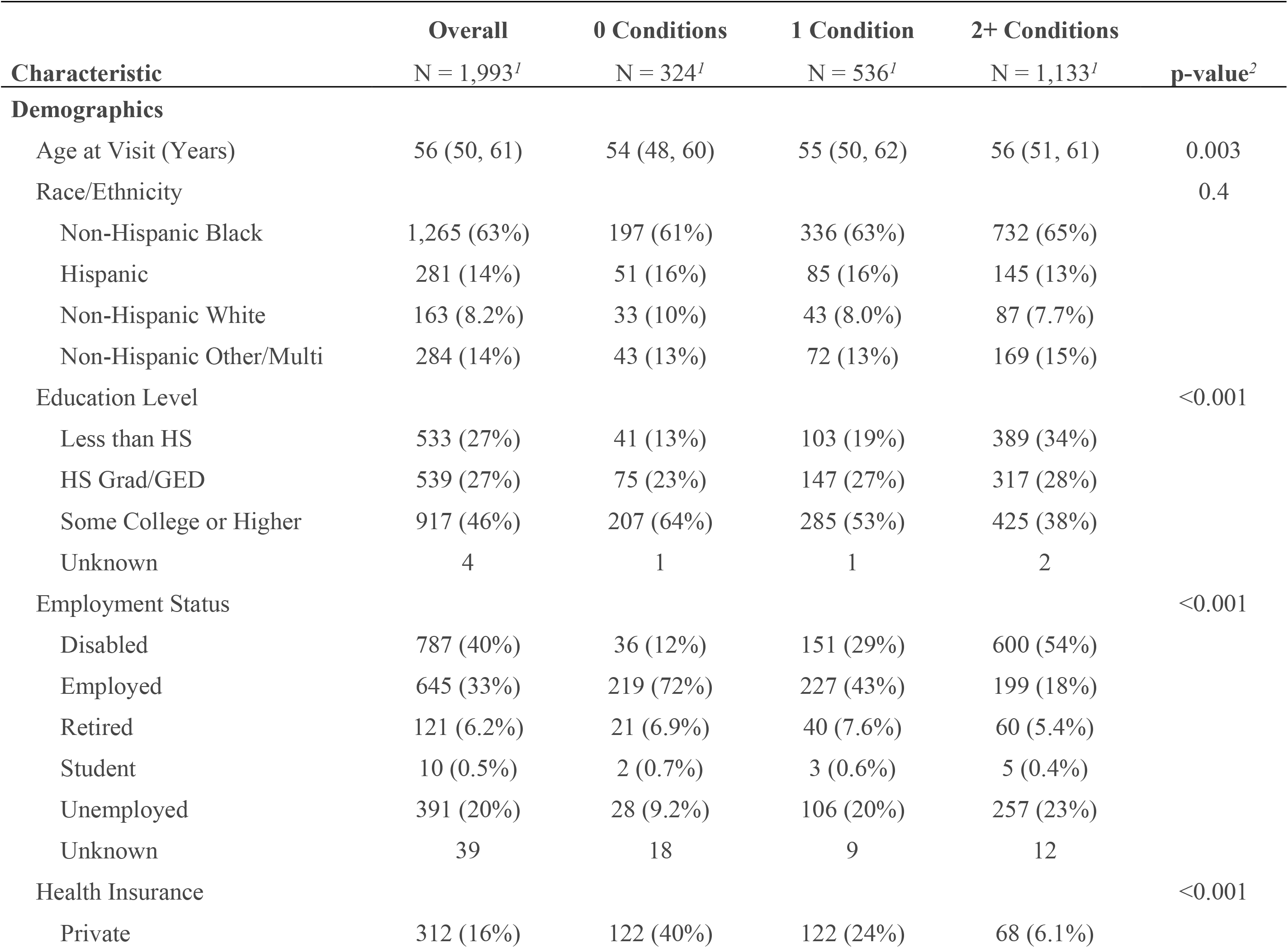
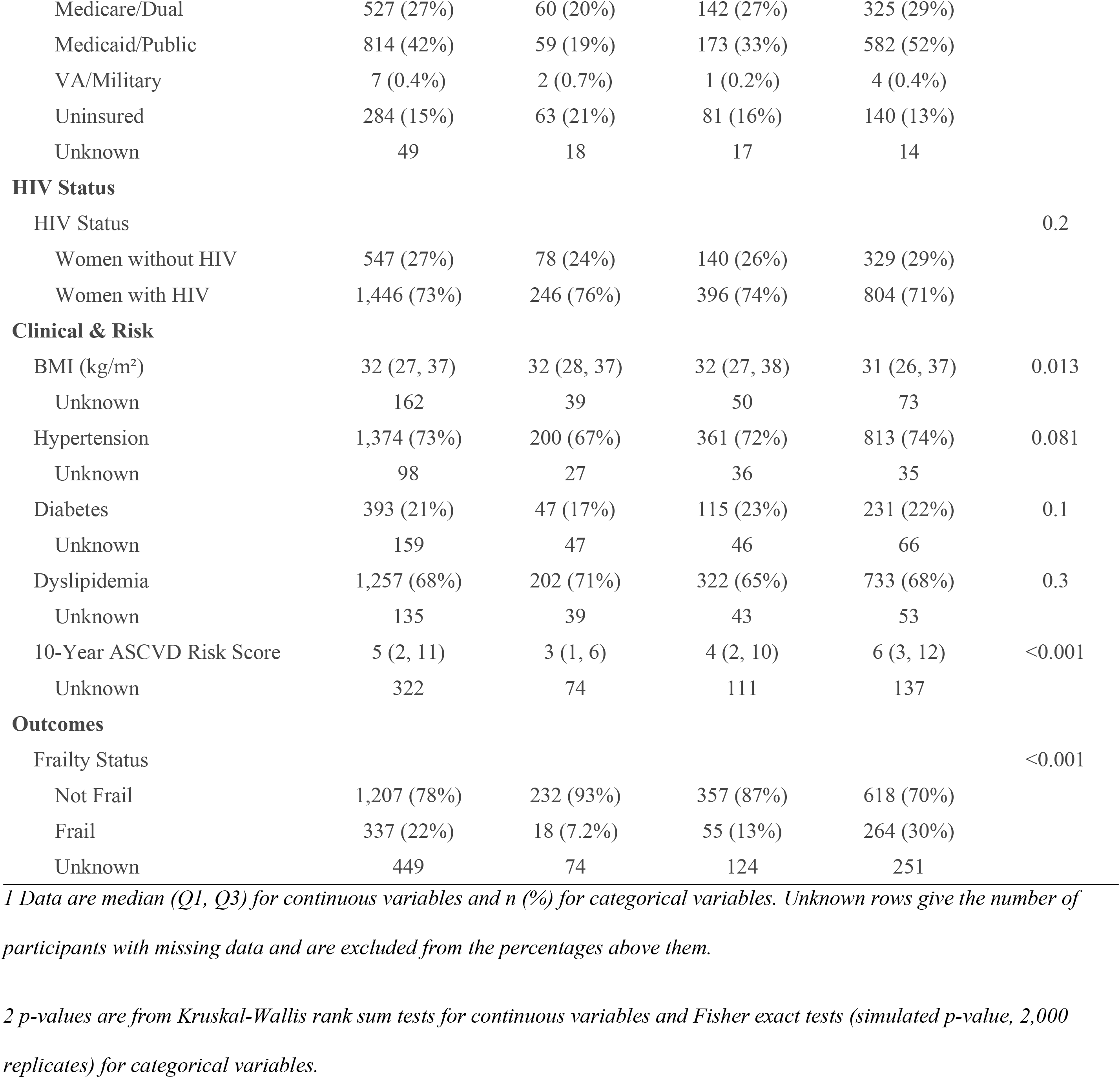
Full sample characteristics by syndemic burden.

|  | **Overall** | **0 Conditions** | **1 Condition** | **2+ Conditions** |  |
|---|---|---|---|---|---|
| **Characteristic** | N = 1,993<sup>*1*</sup> | N = 324<sup>*1*</sup> | N = 536<sup>*1*</sup> | N = 1,133<sup>*1*</sup> | **p-value**<sup>*2*</sup> |
| **Demographics** |  |  |  |  |  |
| Age at Visit (Years) | 56 (50, 61) | 54 (48, 60) | 55 (50, 62) | 56 (51, 61) | 0.003 |
| Race/Ethnicity |  |  |  |  | 0.4 |
| Non-Hispanic Black | 1,265 (63%) | 197 (61%) | 336 (63%) | 732 (65%) |  |
| Hispanic | 281 (14%) | 51 (16%) | 85 (16%) | 145 (13%) |  |
| Non-Hispanic White | 163 (8.2%) | 33 (10%) | 43 (8.0%) | 87 (7.7%) |  |
| Non-Hispanic Other/Multi | 284 (14%) | 43 (13%) | 72 (13%) | 169 (15%) |  |
| Education Level |  |  |  |  | <0.001 |
| Less than HS | 533 (27%) | 41 (13%) | 103 (19%) | 389 (34%) |  |
| HS Grad/GED | 539 (27%) | 75 (23%) | 147 (27%) | 317 (28%) |  |
| Some College or Higher | 917 (46%) | 207 (64%) | 285 (53%) | 425 (38%) |  |
| Unknown | 4 | 1 | 1 | 2 |  |
| Employment Status |  |  |  |  | <0.001 |
| Disabled | 787 (40%) | 36 (12%) | 151 (29%) | 600 (54%) |  |
| Employed | 645 (33%) | 219 (72%) | 227 (43%) | 199 (18%) |  |
| Retired | 121 (6.2%) | 21 (6.9%) | 40 (7.6%) | 60 (5.4%) |  |
| Student | 10 (0.5%) | 2 (0.7%) | 3 (0.6%) | 5 (0.4%) |  |
| Unemployed | 391 (20%) | 28 (9.2%) | 106 (20%) | 257 (23%) |  |
| Unknown | 39 | 18 | 9 | 12 |  |
| Health Insurance |  |  |  |  | <0.001 |
| Private | 312 (16%) | 122 (40%) | 122 (24%) | 68 (6.1%) |  |
| Medicare/Dual | 527 (27%) | 60 (20%) | 142 (27%) | 325 (29%) |  |
| Medicaid/Public | 814 (42%) | 59 (19%) | 173 (33%) | 582 (52%) |  |
| VA/Military | 7 (0.4%) | 2 (0.7%) | 1 (0.2%) | 4 (0.4%) |  |
| Uninsured | 284 (15%) | 63 (21%) | 81 (16%) | 140 (13%) |  |
| Unknown | 49 | 18 | 17 | 14 |  |
| **HIV Status** |  |  |  |  |  |
| HIV Status |  |  |  |  | 0.2 |
| Women without HIV | 547 (27%) | 78 (24%) | 140 (26%) | 329 (29%) |  |
| Women with HIV | 1,446 (73%) | 246 (76%) | 396 (74%) | 804 (71%) |  |
| **Clinical & Risk** |  |  |  |  |  |
| BMI (kg/m²) | 32 (27, 37) | 32 (28, 37) | 32 (27, 38) | 31 (26, 37) | 0.013 |
| Unknown | 162 | 39 | 50 | 73 |  |
| Hypertension | 1,374 (73%) | 200 (67%) | 361 (72%) | 813 (74%) | 0.081 |
| Unknown | 98 | 27 | 36 | 35 |  |
| Diabetes | 393 (21%) | 47 (17%) | 115 (23%) | 231 (22%) | 0.1 |
| Unknown | 159 | 47 | 46 | 66 |  |
| Dyslipidemia | 1,257 (68%) | 202 (71%) | 322 (65%) | 733 (68%) | 0.3 |
| Unknown | 135 | 39 | 43 | 53 |  |
| 10-Year ASCVD Risk Score | 5 (2, 11) | 3 (1, 6) | 4 (2, 10) | 6 (3, 12) | <0.001 |
| Unknown | 322 | 74 | 111 | 137 |  |
| **Outcomes** |  |  |  |  |  |
| Frailty Status |  |  |  |  | <0.001 |
| Not Frail | 1,207 (78%) | 232 (93%) | 357 (87%) | 618 (70%) |  |
| Frail | 337 (22%) | 18 (7.2%) | 55 (13%) | 264 (30%) |  |
| Unknown | 449 | 74 | 124 | 251 |  |

*1 Data are median (Q1, Q3) for continuous variables and n (%) for categorical variables. Unknown rows give the number of participants with missing data and are excluded from the percentages above them.*
*2 p-values are from Kruskal-Wallis rank sum tests for continuous variables and Fisher exact tests (simulated p-value, 2,000 replicates) for categorical variables.*

Baseline characteristics among baseline virally suppressed WLWH are presented in Table 3. In the suppressed WLWH sub-sample (N=1,063), median age was 56 (50, 61), and the sub-sample included 62% non-Hispanic Black (661/1,063), 15% Hispanic (156/1,063), 8.8% non-Hispanic White (94/1,063), and 14% non-Hispanic other/multi (152/1,063) (Table 3). Syndemic burden remained high, with 54% classified as having ≥2 syndemic conditions (579/1,063), 28% with 1 condition (301/1,063), and 17% with no syndemic conditions (183/1,063) (Table 3). Patterns by syndemic burden were similar to the full sample, including strong gradients across education (p<0.001), employment (p<0.001), insurance (p<0.001), baseline ASCVD risk (p<0.001), and frailty (p<0.001; Table 3). Hypertension prevalence increased with syndemic burden (66% in 0 conditions versus 76% in ≥2 conditions; p=0.009; Table 3).

**Table 3.**
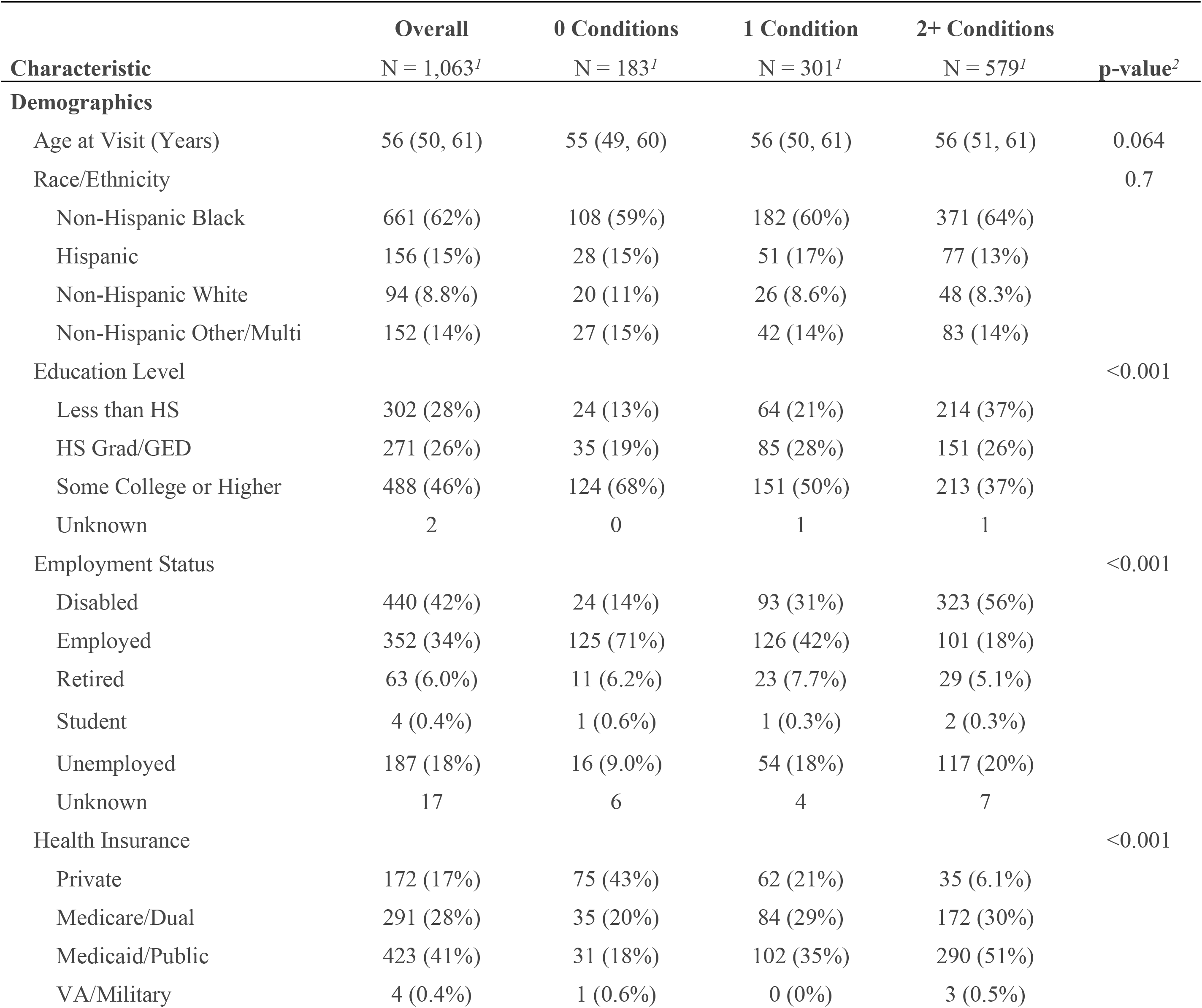
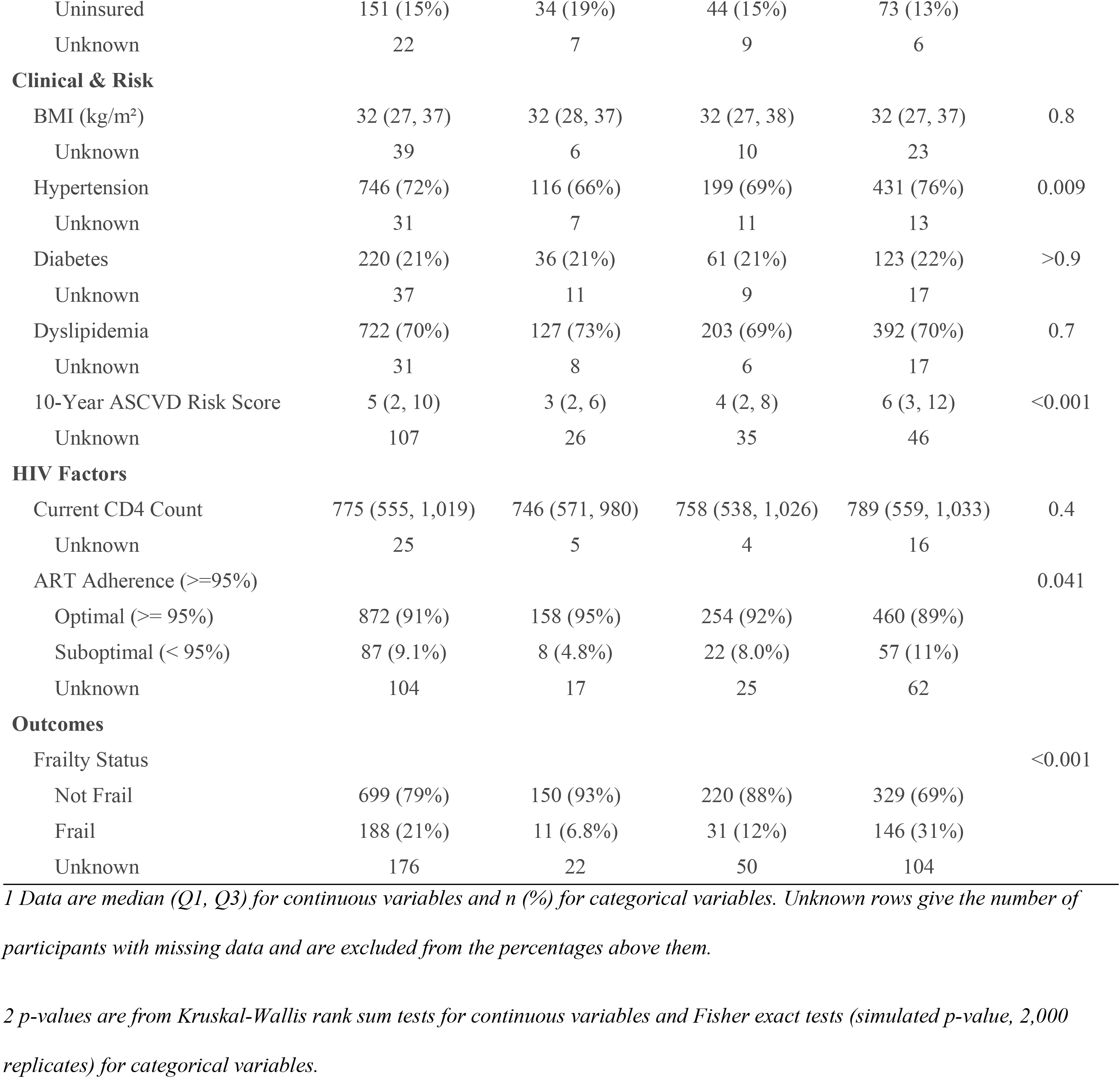
Virally suppressed sub-sample characteristics by syndemic burden.

## Longitudinal mixed-effects models for continuous ASCVD risk

Longitudinal associations in the full analytic sample are presented in Table 4. Continuous ASCVD risk increased over time, with an estimated increase of 0.51 risk-score units per year (β=0.51; 95% CI 0.42, 0.61; p<0.001; Table 4). Higher syndemic score was independently associated with higher ASCVD risk (β=0.40 per 1-point increase; 95% CI 0.01, 0.78; p=0.043; Table 4). Menopause stage was associated with ASCVD risk overall (global p=0.007; Table 4), and postmenopause (natural/surgical) was associated with higher ASCVD risk compared with premenopause (β=1.3; 95% CI 0.39, 2.3; p=0.006; Table 4). The syndemic score-by-menopause stage interaction was not statistically significant overall (global p=0.2; Table 4). HIV status (women with HIV vs women without HIV) was associated with lower ASCVD risk in the specified model (β=-1.4; 95% CI -2.1, -0.64; p<0.001; Table 4).

**Table 4.** Longitudinal association of syndemic burden and menopause stage with 10-year ASCVD risk among women with and without HIV.

| **Characteristic** | **ASCVD Risk** |  |  |
|---|---|---|---|
|  | **Beta** | **95% CI** | **p-value** |
| Time (Years) | 0.51 | 0.42, 0.61 | **<0.001** |
| Syndemic Score | 0.4 | 0.01, 0.78 | **0.043** |
| Menopause Stage |  |  | **0.007** |
| Premenopausal | — | — |  |
| Perimenopause | 0.17 | -0.99, 1.3 | 0.8 |
| Post Menopausal (Nat/Surg) | 1.3 | 0.39, 2.3 | **0.006** |
| HIV Status |  |  | **<0.001** |
| Women without HIV | — | — |  |
| Women with HIV | -1.4 | -2.1, -0.64 | **<0.001** |
| Syndemic Index * Menopause Stage |  |  | 0.2 |
| Syndemic Score * Perimenopause | 0.17 | -0.34, 0.68 | 0.5 |
| Syndemic Score * Post Menopausal (Nat/Surg) | 0.33 | -0.07, 0.73 | 0.1 |

*Linear mixed-effects model with random intercepts for participant; the outcome is the continuous 10-year ASCVD risk score. The terms shown are time in study, syndemic score, menopause stage, the syndemic score by menopause stage interaction, and HIV status. All models were additionally adjusted for the covariates described in Materials and methods: education, employment, insurance, body mass index, hypertension, diabetes, and dyslipidemia. Age and race/ethnicity were not adjusted for because they are direct inputs to the Pooled Cohort Equations and adjusting for them would be circular. ASCVD, atherosclerotic cardiovascular disease; CI, confidence interval.*

Longitudinal associations among baseline virally suppressed WLWH are presented in Table 5. Continuous ASCVD risk increased over time (β=0.59 per year; 95% CI 0.47, 0.71; p<0.001; Table 5). In this sub-sample, the main effect of syndemic score was not statistically significant (β=0.39; 95% CI -0.15, 0.92; p=0.2; Table 5), and menopause stage was not significant overall (global p=0.3; Table 5), although the syndemic score-by-postmenopause interaction suggested an exploratory signal (β=0.50; 95% CI -0.06, 1.1; p=0.079; Table 5).

**Table 5.**
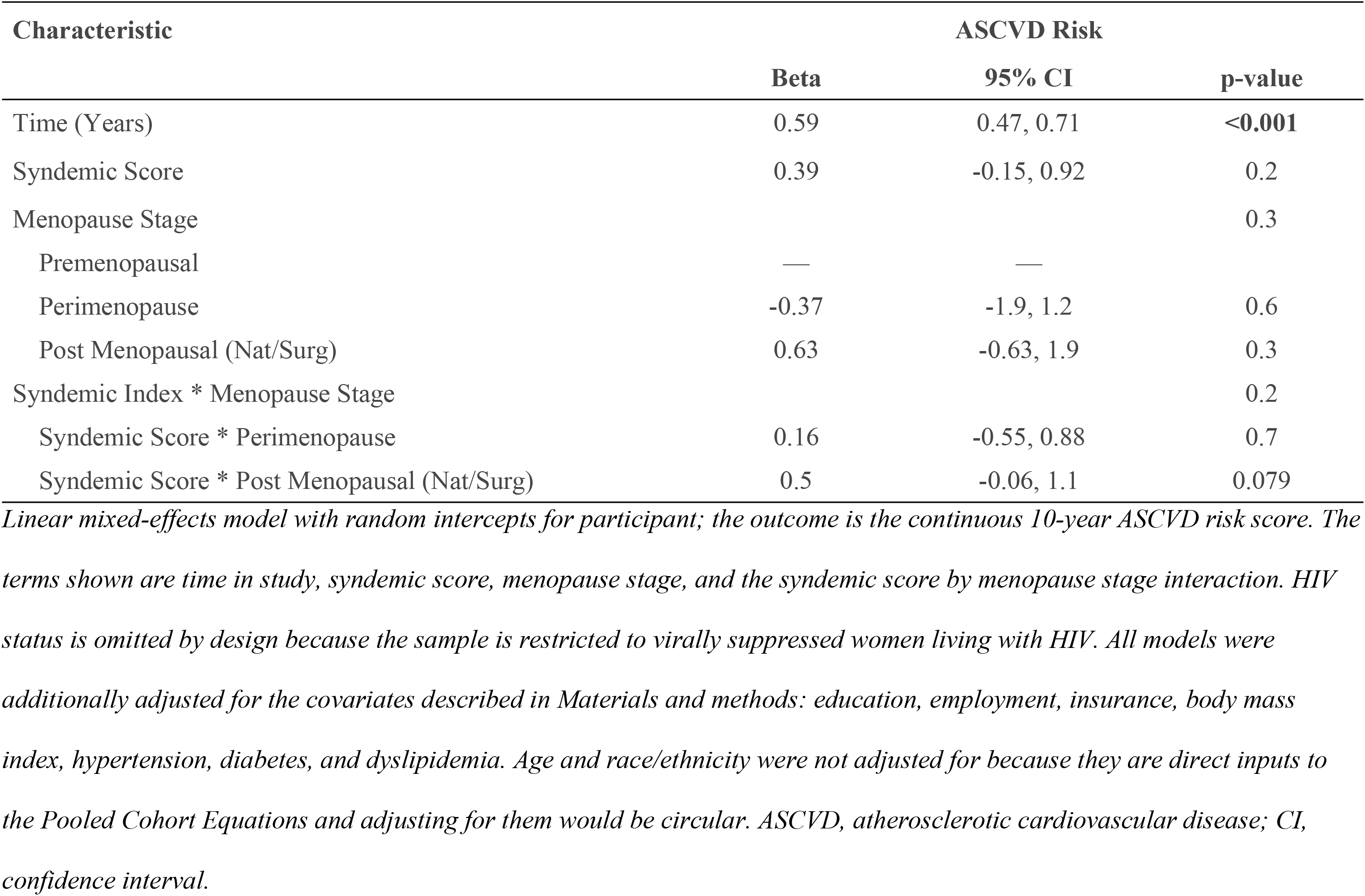
Longitudinal association of syndemic burden and menopause stage with 10-year ASCVD risk among virally suppressed women living with HIV.

## Random forest classifiers predicting high ASCVD risk

Random forest performance and variable importance (VIP) results for the full analytic sample are presented in Fig 2. For prediction of high ASCVD risk (≥7.5%), the model demonstrated moderate discrimination (AUC=0.727) and accuracy 0.654, with sensitivity 0.686, specificity 0.627, PPV 0.613, and NPV 0.699 (Fig 2). Predictors ranked among the most important for high ASCVD risk included FIB-4, postmenopausal status, triglycerides, serum creatinine, BMI, and the syndemic score, with loneliness and dyslipidemia-related indicators also appearing among influential features (Fig 2).

**Fig 2.**
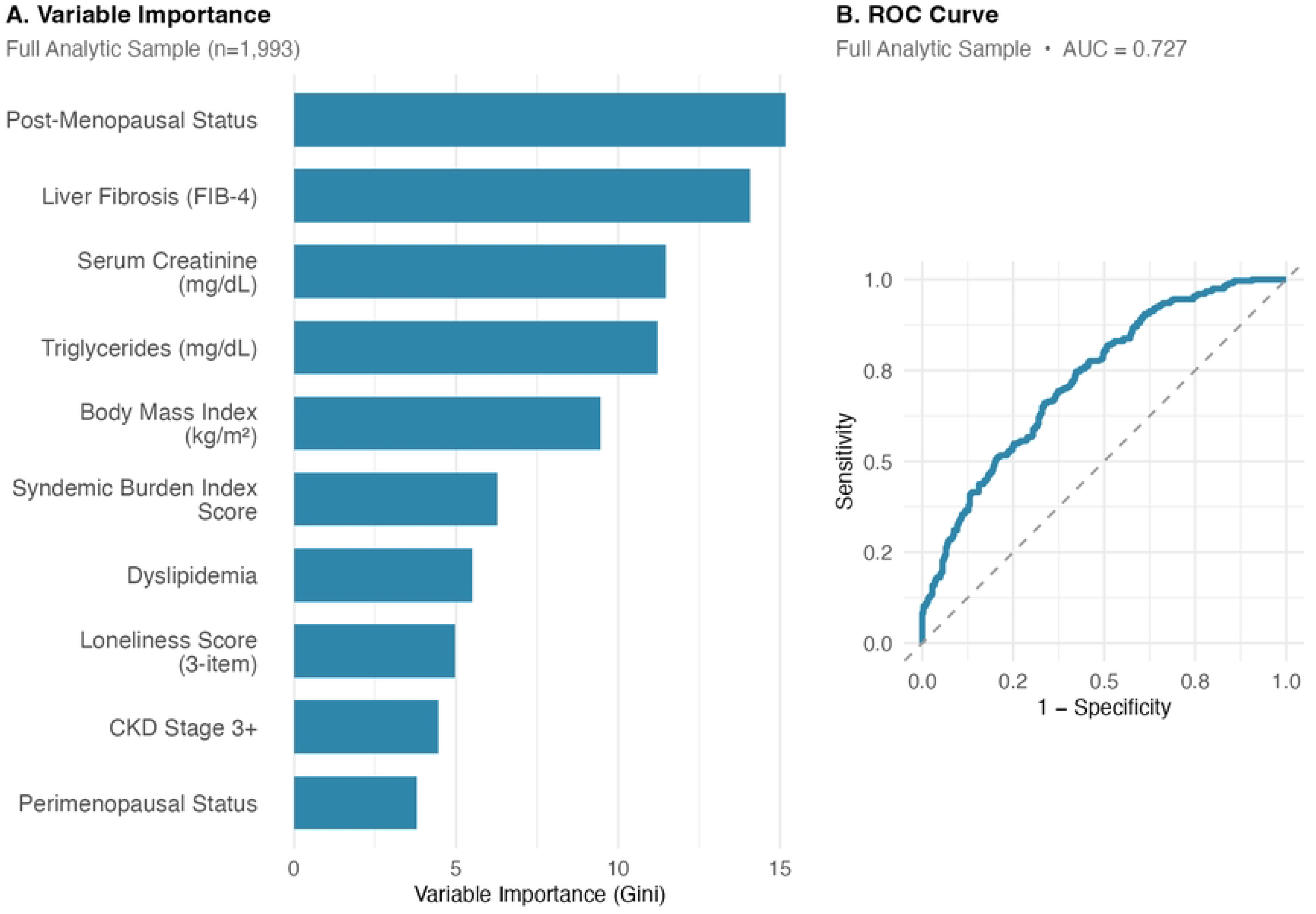
Random forest classifier predicting high 10-year ASCVD risk (≥7.5%) in the full analytic cohort. Full analytic cohort (N=1,993). Panel A shows the 10 most important predictors by Gini impurity-based variable importance; panel B shows the receiver operating characteristic curve (AUC=0.727). The 32-predictor set excluded the Pooled Cohort Equation input variables to avoid circularity. Model settings: 1,000 trees, 5-fold cross-validation, 70/30 train-test split, and downsampling for class imbalance. ASCVD, atherosclerotic cardiovascular disease; AUC, area under the curve; WLWH, women living with HIV.

Random forest results for the virally suppressed WLWH sub-sample are presented in Fig 3. For prediction of high ASCVD risk (≥7.5%), discrimination remained similar (AUC=0.704) with accuracy 0.646, sensitivity 0.740, specificity 0.568, PPV 0.584, and NPV 0.727 (Fig 3). The most important predictors included postmenopausal status, serum creatinine, BMI, hypertension indicators, and syndemic score, with poverty and depression also contributing prominently (Fig 3).

**Fig 3.**
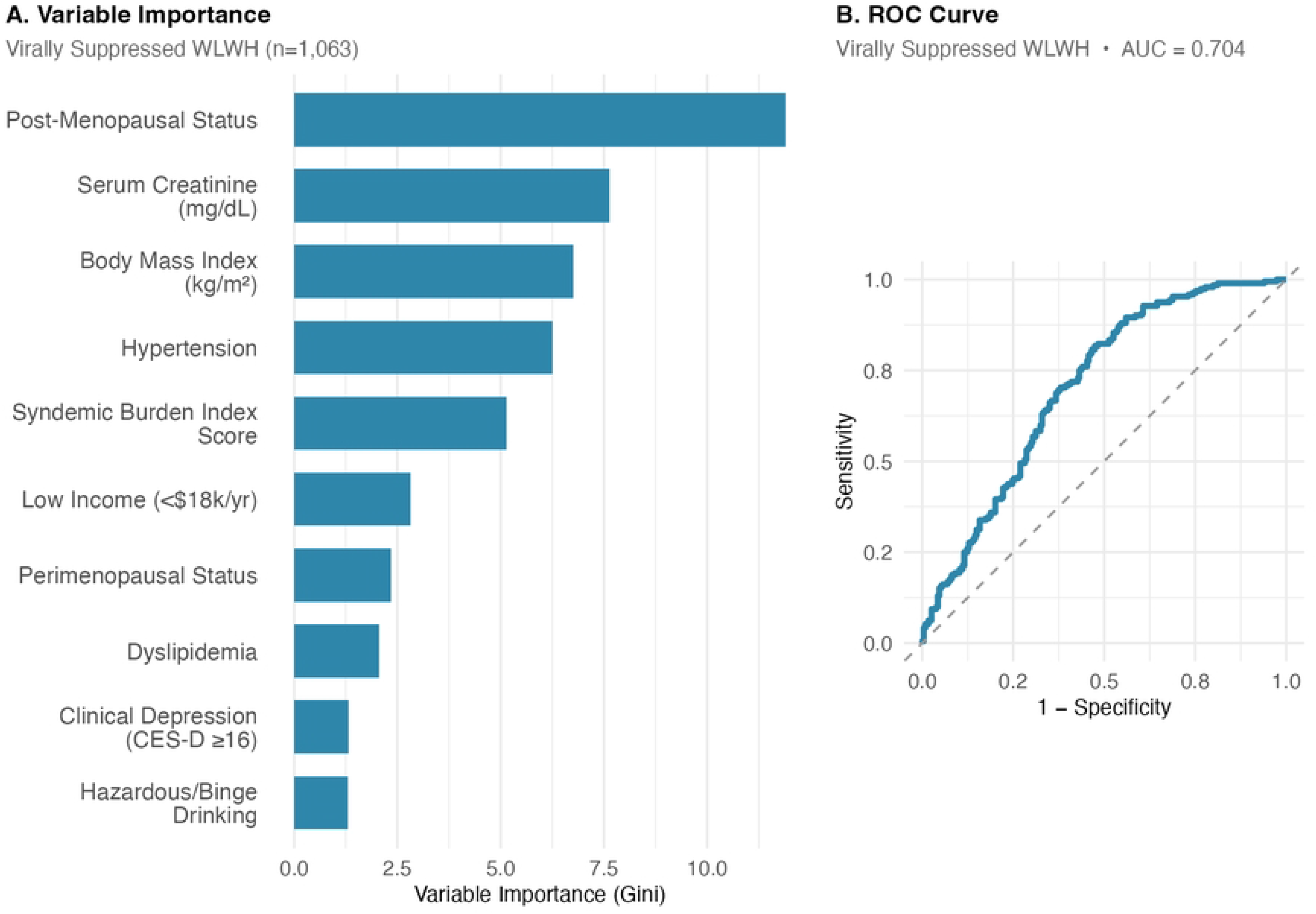
Random forest classifier predicting high 10-year ASCVD risk (≥7.5%) among virally suppressed women living with HIV. Virally suppressed sub-cohort (N=1,063; 4,009 person- visits). Panel A shows the 10 most important predictors by Gini impurity-based variable importance; panel B shows the receiver operating characteristic curve (AUC=0.704). Predictors and model settings are identical to those in Fig 2, except for a 60/40 train-test split. ASCVD, atherosclerotic cardiovascular disease; AUC, area under the curve.

Cross-model synthesis of variable importance is summarized in Fig 4. Across both random forest models (full cohort and virally suppressed sub-cohort predicting high ASCVD risk), serum creatinine, BMI, syndemic score, and postmenopausal status consistently ranked among the top predictors (Fig 4).

**Fig 4.**
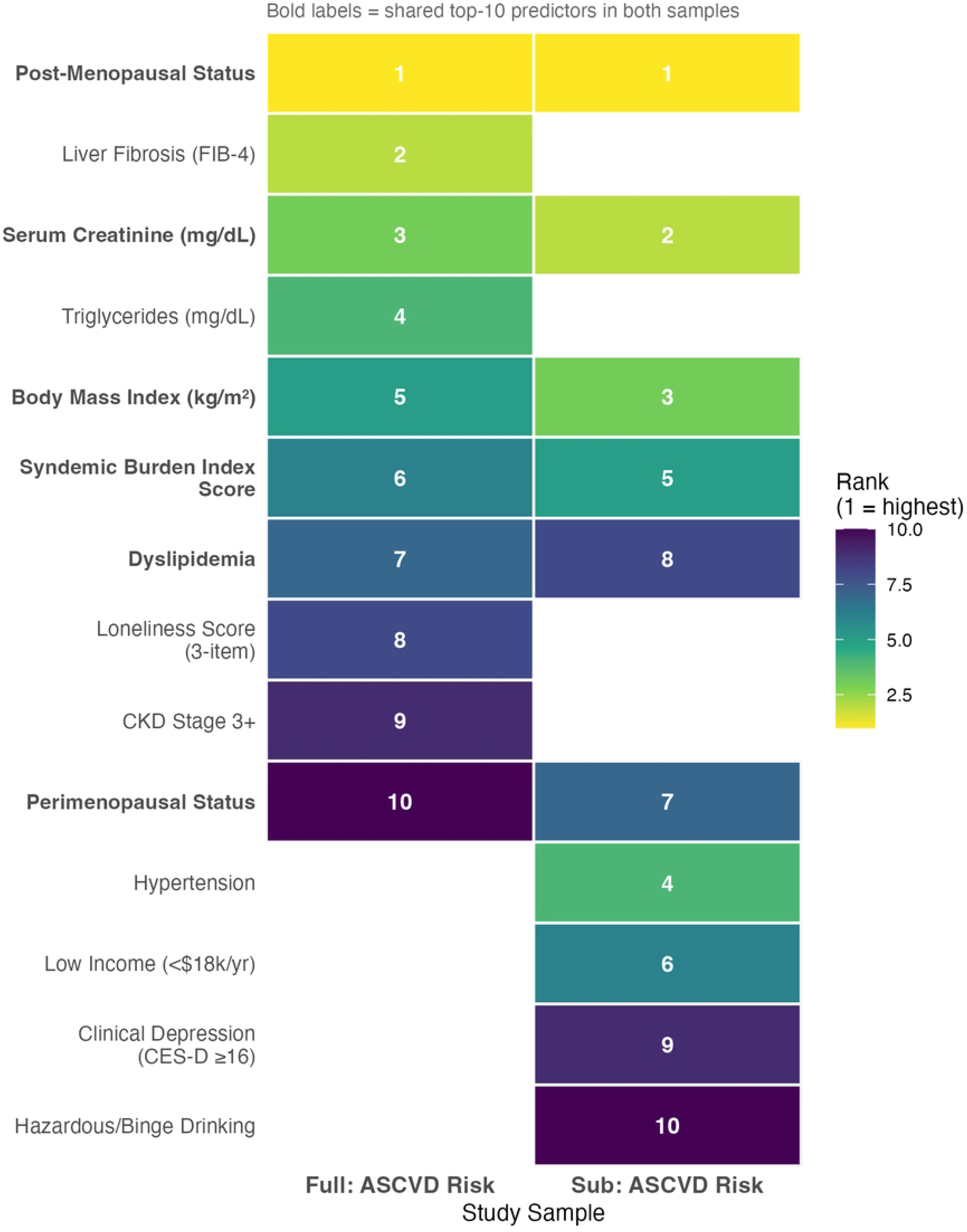
Variable importance ranks compared across the full cohort and the virally suppressed sub-cohort. Rank of each predictor within the top 10 of the two random forest classifiers for high 10-year ASCVD risk (≥7.5%): the full cohort (N=1,993) and the virally suppressed sub-cohort (N=1,063). Rank 1 is the most important predictor; blank cells indicate a predictor outside that model’s top 10. Bold row labels mark predictors shared by both models. Serum creatinine, body mass index, syndemic burden, and postmenopausal status ranked among the top predictors in both samples. ASCVD, atherosclerotic cardiovascular disease.

## Discussion

### Summary of principal findings

In this longitudinal analysis of midlife and older women in the MWCCS, higher syndemic burden and postmenopausal status were each independently associated with higher estimated ASCVD risk in the full analytic sample. Each additional syndemic condition was associated with a statistically significant but modest increase in continuous estimated ASCVD risk, and postmenopausal status was associated with higher estimated ASCVD risk compared with premenopause. The syndemic-by-menopause interaction was not statistically significant overall, though an exploratory signal was observed among virally suppressed WLWH. Random forest classifiers ranked syndemic score, BMI, serum creatinine, and postmenopausal status among the consistent top predictors of high estimated ASCVD risk across both analytic populations.

Machine learning approaches complement traditional regression by capturing potential nonlinear relationships and providing hypothesis-generating variable prioritization for future risk prediction tool development.

## Menopause stage and syndemic burden as independent predictors of estimated ASCVD risk

Importantly, menopause is associated with a narrowing of the traditional sex-based protection against cardiovascular disease. As endogenous estrogen declines during the menopausal transition, estrogen-mediated vasodilatory and anti-inflammatory effects are attenuated, and women’s cardiovascular risk profiles converge toward those of age-matched men. This phenomenon is clinically relevant because it situates postmenopausal WLWH - who face both HIV-related immune activation and the hormonal shifts of menopause - at potentially heightened cardiovascular vulnerability. [19,20,21]

The observed association between postmenopausal status and higher estimated ASCVD risk is biologically plausible: the menopause transition involves endocrine changes associated with adverse shifts in lipid profiles, body fat distribution, and vascular function - all upstream determinants of ASCVD risk [8,19]. These findings are consistent with evidence from the REPRIEVE trial that cardiovascular risk in WLWH is shaped by reproductive aging and immune activation beyond what chronological age alone captures [4,11].

Syndemic burden plausibly contributes to ASCVD risk through chronic stress biology, material hardship constraining preventive care, and barriers to health behaviors [5]. These results extend prior work linking behavioral and psychosocial syndemics to systemic inflammation and cardiometabolic risk [7,22]. The consistent ranking of syndemic score among top predictors in both random forest models reinforces its value as a summary index of modifiable vulnerability.

Although the syndemic-by-postmenopause interaction was not significant in the full sample, the exploratory signal in the virally suppressed sub-cohort (p=0.079) suggests that postmenopausal WLWH with higher syndemic burden may warrant further evaluation. Larger samples or longer follow-up are needed to evaluate this possible amplification effect.

These findings have clinical relevance in the context of updated cardiovascular prevention guidelines. The 2026 AHA/ACC cholesterol management guidelines recommend LDL-lowering therapy for adults aged 40-75 years with HIV irrespective of LDL-C level. Our finding that syndemic burden was associated with higher estimated ASCVD risk, after accounting for HIV status and selected clinical and social covariates while avoiding circular adjustment for PCE-embedded variables, suggests that syndemic screening may help identify WLWH who could benefit from intensified cardiovascular prevention efforts. [23]

## Strengths and limitations

MWCCS provides a large, well-characterized cohort enabling analyses that integrate reproductive aging and syndemic frameworks within a longitudinal design. The analytic approach combined mixed-effects inference and prediction modeling, explicitly addressing circularity concerns by excluding PCE components from machine learning predictors.

Several limitations require consideration. First, ASCVD risk was a computed estimator rather than an observed event outcome, and PCE-based risk estimation has recognized limitations in HIV populations, including evidence of underestimation and sex differences in calibration; associations with the risk score should not be interpreted as direct evidence of proportional changes in adjudicated event incidence [10]. Because the PCE incorporates age, the estimated time effect partly reflects aging embedded in the risk equation as well as changes in measured risk factors over follow-up. Second, the PREVENT equation [24] - a more contemporary risk tool that incorporates kidney function and a social deprivation index - was available from the DACC at the V1051 visit only, which did not support the full longitudinal analysis spanning V101 through V1051; future work should validate these findings using PREVENT once data become available across all visits. Third, individual PCE components were intentionally excluded from the random forest predictor set; however, current tobacco use is included within the composite syndemic score, so the syndemic score is not fully independent of all PCE inputs. Physical inactivity is not a PCE component but is correlated with cardiometabolic risk, and both features may influence random forest variable importance for estimated ASCVD risk models.

Because impurity-based Gini importance can favor continuous or high-cardinality predictors, variable-importance rankings should be interpreted as descriptive prioritization; future work should examine permutation importance and external validation. Fourth, syndemic burden components were equally weighted (each scored 0/1); sensitivity analyses with differential weighting or latent class approaches were not performed and may yield different findings.

Additionally, because syndemic components were measured cross-sectionally at each visit, the temporal ordering of syndemic burden relative to changes in ASCVD risk cannot be established, and causal inference is not supported. Fifth, menopause staging was based on self-reported menses and documented surgical history; menopause status was treated as a time-varying covariate updated at each study visit based on self-reported menses and documented surgical history; cases with apparent reversal from postmenopausal to perimenopausal staging were retained as reported and imputed using polytomous regression in the multiple imputation step.

Finally, non-adjudicated CVD events were not included as an outcome in this analysis, as MWCCS event adjudication is ongoing; associations between syndemic burden and incident cardiovascular events will be examined in future work once adjudicated data are available.

## Conclusion

Among midlife and older women in the MWCCS, syndemic burden and postmenopausal status were each independently associated with higher longitudinal estimated ASCVD risk. Random forest classifiers demonstrated moderate discrimination for high estimated ASCVD risk, with syndemic score, BMI, serum creatinine, and postmenopausal status consistently ranked among the top predictors. These findings support integrating both reproductive aging stage and syndemic burden into cardiovascular risk assessment strategies for women aging with and without HIV, and support the need for syndemic-informed prevention approaches in this population.

## Data Availability

Data cannot be shared publicly because of participant confidentiality protections. MWCCS data are available to qualified researchers through the NIH-supported MWCCS Data Analysis and Coordination Center (DACC), which reviews data requests rather than the authors. Requests may be submitted at https://mwccs.org/data-access in accordance with NIH data-sharing policies. The analytic code implementing the mixed-effects models and random forest classifiers is available on reasonable request from the corresponding author and will be deposited in a public repository on publication.

## Acknowledgments

The authors gratefully acknowledge the contributions of the study participants and dedication of the staff at the MWCCS sites.

The contents of this publication are solely the responsibility of the authors and do not represent the official views of the National Institutes of Health (NIH). MWCCS (Principal Investigators): Atlanta CRS (Ighovwerha Ofotokun, Anandi Sheth, and Gina Wingood), U01-HL146241; Baltimore CRS (Todd Brown and Joseph Margolick), U01-HL146201; Bronx CRS (Kathryn Anastos, David Hanna, and Anjali Sharma), U01-HL146204; Brooklyn CRS (Deborah Gustafson and Tracey Wilson), U01-HL146202; Data Analysis and Coordination Center (Gypsyamber D’Souza, Stephen Gange and Elizabeth F. Topper), U01-HL146193; Chicago- Cook County CRS (Mardge Cohen, Audrey French, and Ryan Ross), U01-HL146245; Chicago- Northwestern CRS (Steven Wolinsky, Frank Palella, and Valentina Stosor), U01-HL146240; Northern California CRS (Bradley Aouizerat, Jennifer Price, and Phyllis Tien), U01-HL146242; Los Angeles CRS (Roger Detels and Matthew Mimiaga), U01-HL146333; Metropolitan Washington CRS (Seble Kassaye and Daniel Merenstein), U01-HL146205; Miami CRS (Maria Alcaide, Claudia Martinez, and Deborah Jones), U01-HL146203; Pittsburgh CRS (Jeremy Martinson and Charles Rinaldo), U01-HL146208; UAB-MS CRS (Mirjam-Colette Kempf, James B. Brock, Emily Levitan, and Deborah Konkle-Parker), U01-HL146192; UNC CRS (M. Bradley Drummond and Michelle Floris-Moore), U01-HL146194. The MWCCS is funded primarily by the National Heart, Lung, and Blood Institute (NHLBI), with additional co-funding from the Eunice Kennedy Shriver National Institute of Child Health & Human Development (NICHD), National Institute on Aging (NIA), National Institute of Dental & Craniofacial Research (NIDCR), National Institute of Allergy and Infectious Diseases (NIAID), National Institute of Neurological Disorders and Stroke (NINDS), National Institute of Mental Health (NIMH), National Institute on Drug Abuse (NIDA), National Institute of Nursing Research (NINR), National Cancer Institute (NCI), National Institute on Alcohol Abuse and Alcoholism (NIAAA), National Institute on Deafness and Other Communication Disorders (NIDCD), National Institute of Diabetes and Digestive and Kidney Diseases (NIDDK), National Institute on Minority Health and Health Disparities (NIMHD), and in coordination and alignment with the research priorities of the National Institutes of Health, Office of AIDS Research (OAR).

MWCCS data collection is also supported by UL1-TR000004 (UCSF CTSA), UL1-TR003098 (JHU ICTR), UL1-TR001881 (UCLA CTSI), P30-AI-050409 (Atlanta CFAR), P30-AI-073961 (Miami CFAR), P30-AI-050410 (UNC CFAR), P30-AI-027767 (UAB CFAR), P30-AI-124414 (ERC-CFAR), P30-MH-116867 (Miami CHARM), UL1-TR001409 (DC CTSA), KL2- TR001432 (DC CTSA), and TL1-TR001431 (DC CTSA).

## Data availability

MWCCS data are available to qualified researchers through the NIH-supported MWCCS Data Analysis and Coordination Center. Data requests may be submitted at https://mwccs.org/data- access in accordance with NIH data-sharing policies. Requests are reviewed by the Data Analysis and Coordination Center rather than by the authors. Individual-level data cannot be publicly posted because of participant confidentiality protections.

Analyses were conducted in R (version 4.5.2) using the tidymodels, lme4, and mice packages. The analytic code implementing the mixed-effects models and the random forest classifiers is available on reasonable request from the corresponding author and will be deposited in a public repository on publication.

All survey instruments, including the Center for Epidemiologic Studies Depression (CES-D) scale, the AUDIT-C, and the syndemic component definitions, are documented in the MWCCS data manuals and can be obtained through the MWCCS website or from the Data Analysis and Coordination Center.

